# Health conditions and RSV-related Pediatric Intensive Care Unit admissions in children during their second RSV season

**DOI:** 10.64898/2026.06.26.26356705

**Authors:** Regina M. Simeone, Laura D. Zambrano, Margaret M. Newhams, Amanda B. Payne, Amber O. Orzel-Lockwood, Natasha B. Halasa, Jemima Calixte, Aline B. Maddux, Kathleen Chiotos, Satoshi Kamidani, Hillary Crandall, Danielle M. Zerr, Melissa A. Cameron, Shira J. Gertz, Bria M. Coates, Kelly N. Michelson, Jennifer E. Schuster, Ryan A. Nofziger, Jigar C. Chauhan, Mia Maamari, Steven L. Shein, Michele Kong, Janet R. Hume, Lora M. Martin, Judith A. Guzman-Cottrill, Samina S. Bhumbra, Katherine Irby, Mary Allen Staat, Tamara T. Bradford, Kari Wellnitz, Melissa S. Stockwell, Matt Zinter, Stephanie P. Schwartz, Saul Hymes, Emily R. Levy, Austin Biggs, Katherine Lindsey, Angela P. Campbell, Adrienne G. Randolph, the Overcoming RSV investigators

## Abstract

**Importance:** Respiratory syncytial virus (RSV) hospitalization rates are highest among children <2 years of age. RSV immunization with infant monoclonal antibody or maternal vaccine is recommended to protect all U.S. infants in their first RSV season. For certain high-risk children aged 8−19 months entering their second RSV season, the monoclonal antibody nirsevimab is recommended. Little is known regarding preexisting health conditions as risk factors for RSV-associated respiratory failure in children during their second season.

**Objectives:** To describe children admitted to the pediatric intensive care unit (PICU) for RSV during their second RSV season by preexisting health conditions, and to compare demographic and clinical characteristics across groups.

**Design, Setting, and Participants:** Surveillance registry of children 8-<24 months old admitted to the PICU in 30 pediatric hospitals in the 2023-2024/2024-2025 RSV seasons. All children had an RSV-positive respiratory sample and received respiratory support with high flow nasal cannula, noninvasive ventilation, or invasive mechanical ventilation (IMV).

**Exposure:** Preexisting health conditions potentially increasing risk of severe RSV disease.

**Main Outcomes and Measures:** Patients were classified into four mutually exclusive groups by preexisting health conditions: 1) U.S. nirsevimab eligible criteria, 2) other identified RSV risk conditions (with some evidence of increased risk for severe RSV), 3) other preexisting conditions, and 4) no preexisting conditions. Patient demographic characteristics and level of respiratory support received were compared.

**Results:** Among 574 children: 47 (8.2%) had U.S. nirsevimab eligibility criteria, 76 (13.2%) had other RSV risk conditions, 96 (16.7%) had other preexisting conditions, and 355 (61.8%) had none. A higher proportion of children with nirsevimab eligibility factors (40.4%) than those with other identified RSV risk conditions (17.1%) required IMV, which was higher than other (10.4%) or no (5.9%) preexisting health conditions (p_trend_<0.001).

**Conclusions and Relevance:** Approximately 20% of children admitted to the PICU with severe RSV were in the defined groups that met U.S. nirsevimab-eligibility criteria or that had an identified RSV risk condition associated with known risk for severe RSV. A considerable proportion of both groups of children required IMV for respiratory support. These findings may help inform future deliberations regarding U.S. second season nirsevimab-eligibility recommendations.

**Key Points:** *Question:* What preexisting health conditions are present in children aged 8–<24 months admitted to the pediatric intensive care unit for severe RSV disease in their second RSV season and how do clinical outcomes differ?

*Findings:* Among 574 children, 8% were eligible for nirsevimab in the U.S. and 13% had other preexisting conditions with known risk for severe RSV. Children with preexisting conditions, particularly those eligible for nirsevimab, were more likely to require invasive mechanical ventilation than those without.

*Meaning:* These findings may inform future deliberations regarding which children in their second RSV season might benefit from RSV preventive products.

## Introduction

Respiratory syncytial virus (RSV) is an important cause of acute lower respiratory tract infections in children under the age of two years.^1,2^ Among infants hospitalized for RSV, including among those who require intensive care for RSV, most are born at term and are previously healthy with no preexisting health conditions.^3,4^ In contrast to infants, fewer data are available to inform knowledge of clinical characteristics of children who experience severe RSV disease during their second RSV season.^5,6^ While rates of RSV-associated hospitalizations in United States (U.S.) children are highest for infants in the first months of life, they remain elevated in young children aged 12–23 months compared with children 24–59 months.^1,3,7^ Children with RSV can experience RSV bronchiolitis, pneumonia, the need for respiratory support, pediatric intensive care unit (PICU) admission, and death.^8,9^

Until 2023, the only approved product to protect against RSV in a child’s first or second RSV season was palivizumab, a short-acting monoclonal antibody. Palivizumab was a costly intervention, required monthly dosing during RSV season, and was recommended only for infants and young children with specific preexisting health conditions.^5,6,10,11^ In August 2023, the U.S. Advisory Committee on Immunization Practices recommended that all U.S. infants <8 months be protected from severe RSV by either maternal RSV vaccination during pregnancy or by infant receipt of anti-RSV long-acting monoclonal antibody (i.e., nirsevimab, if maternal RSV vaccination was not administered).^5,10^ Further, infants and children aged 8−19 months with specific preexisting health conditions at increased risk for severe disease entering their second RSV season were also recommended to receive nirsevimab.^5,10,12^ Second season recommendations were based on previous palivizumab eligibility criteria, cost effectiveness, and data demonstrating that American Indian and Alaska Native children experienced high rates of RSV disease.^5,6^ Additional risk factors were not added due to insufficient epidemiologic data.^5^ Palivizumab is not routinely recommended given its expense, short duration of protection, and monthly administration challenges.^10 13,14^

The second season U.S. nirsevimab eligibility criteria include: 1) chronic lung disease (CLD) of prematurity requiring medical support (chronic corticosteroid therapy, diuretic therapy, or supplemental oxygen) during the 6-month period before the start of the second RSV season, 2) severe immunocompromise, 3) cystic fibrosis with either manifestations of severe lung disease or weight-for-length <10^th^ percentile, or 4) being American Indian or Alaska Native (AI/AN). Data are needed to better understand which infants and young children entering their second RSV season may be at increased risk of severe RSV disease but do not meet one of the current U.S. eligibility criteria, and who might benefit from nirsevimab or a similar, future licensed preventive product. In addition to the current U.S. nirsevimab eligibility factors, other identified conditions that increase RSV risk include hemodynamically significant cardiac disease, neuromuscular disorders, congenital airway or pulmonary abnormalities, prematurity, trisomy 21, and other genetic disorders, which have been included as eligibility criteria for some other countries.^1,6,15–17^ The objective of this investigation was to describe infants and young children aged 8 to <24 months in their second RSV season who were admitted to the PICU for severe RSV by classifying them into four mutually exclusive groups by their preexisting health conditions status or AI/AN status: 1) U.S. nirsevimab eligible criteria, 2) other identified RSV risk conditions, 3) other preexisting conditions, and 4) no preexisting conditions. We then compared the demographics and clinical characteristics across preexisting health conditions groups.

## Methods

The CDC *Overcoming RSV* network conducts hospital-based surveillance for RSV-associated PICU admissions among children aged <24 months experiencing their first and second RSV seasons.^18^ This analysis was limited to children entering their second RSV season (8−19 months of age on October 1^st^ of the 2023−2024 or 2024−2025 RSV season, depending on the year of enrollment) and <24 months of age at the time of hospitalization across 30 hospitals in 25 U.S. states in a critical care registry between October 30, 2023 to April 15, 2024 (study year 1) or October 1, 2024 to April 15, 2025 (study year 2). All children had a positive RSV test performed in a healthcare setting within 72 hours of admission. Analytic inclusion required 1) admission to the PICU for ≥24 hours and 2) receipt of high flow nasal cannula, noninvasive ventilation (continuous positive airway pressure [CPAP] or bilevel positive airway pressure [BiPAP]), and/or invasive mechanical ventilation during the PICU stay. Receipt of respiratory support was required as confirmation of RSV disease severity.

Children were categorized into four mutually exclusive eligibility status and preexisting health conditions groups: 1) “U.S. nirsevimab eligible criteria” (including AI/AN status); 2) “other identified RSV risk conditions”; 3) “other conditions” (e.g., asthma/reactive airway disease); and 4) “no conditions”. Children were assigned to one mutually exclusive category using a hierarchical approach. Those with any U.S. nirsevimab eligibility criterion were classified as U.S nirsevimab eligible, even if they also had other identified RSV risk conditions or other conditions. Among children without U.S. nirsevimab eligible criteria, those with other identified RSV-risk conditions were classified in that group, even if they also had other conditions not known to be associated with increased risk for severe RSV disease. Nirsevimab eligibility was determined in accordance with current U.S. recommendations.^5^ Other identified RSV risk conditions were clinically significant health conditions determined *a priori* based on limited published evidence of increased risk for severe RSV-associated disease among children in their second RSV season and on second season nirsevimab recommendations from countries other than the United States (Supp. Table 1).^1,5,6,15–17^

**Table 1.**
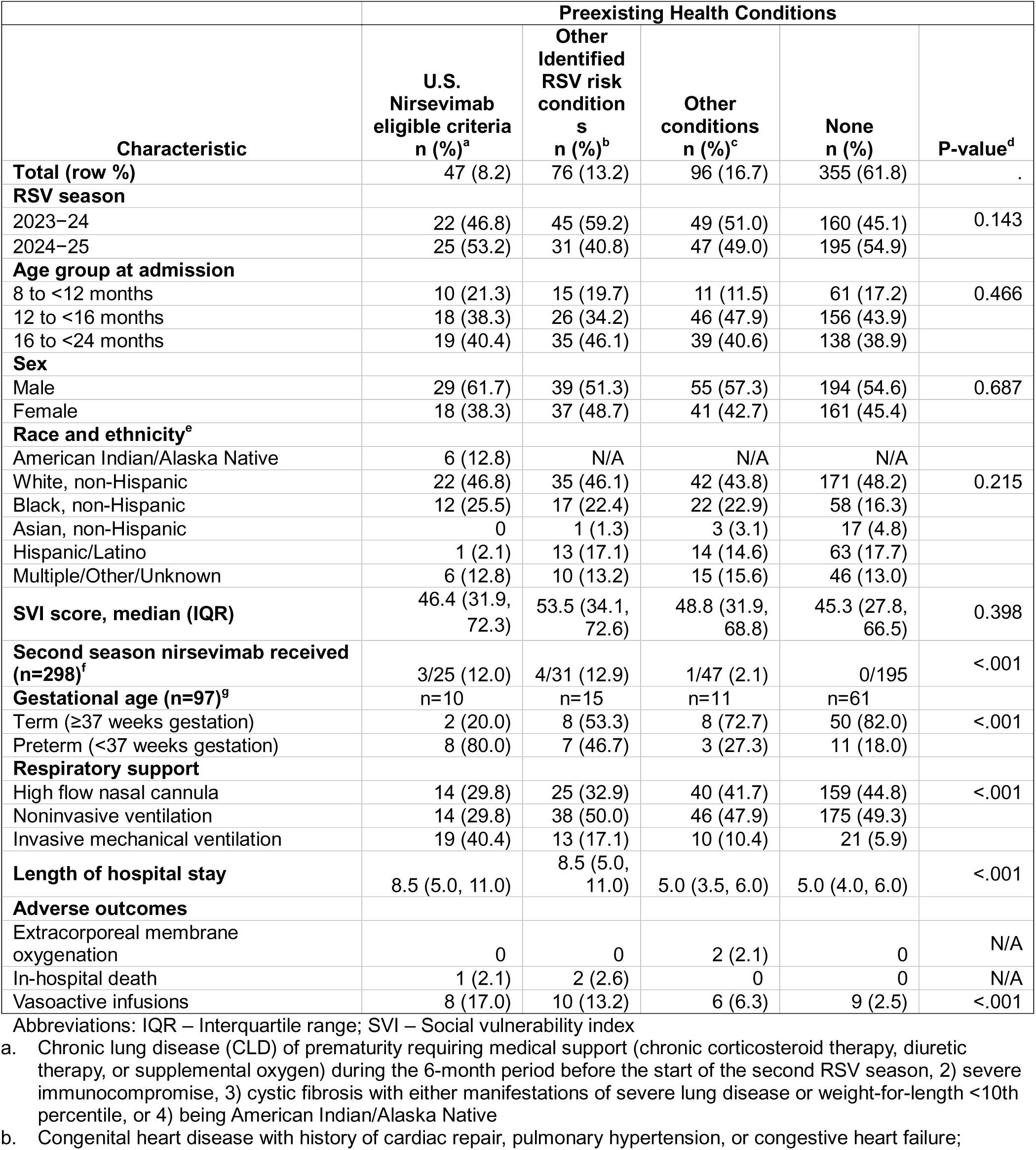
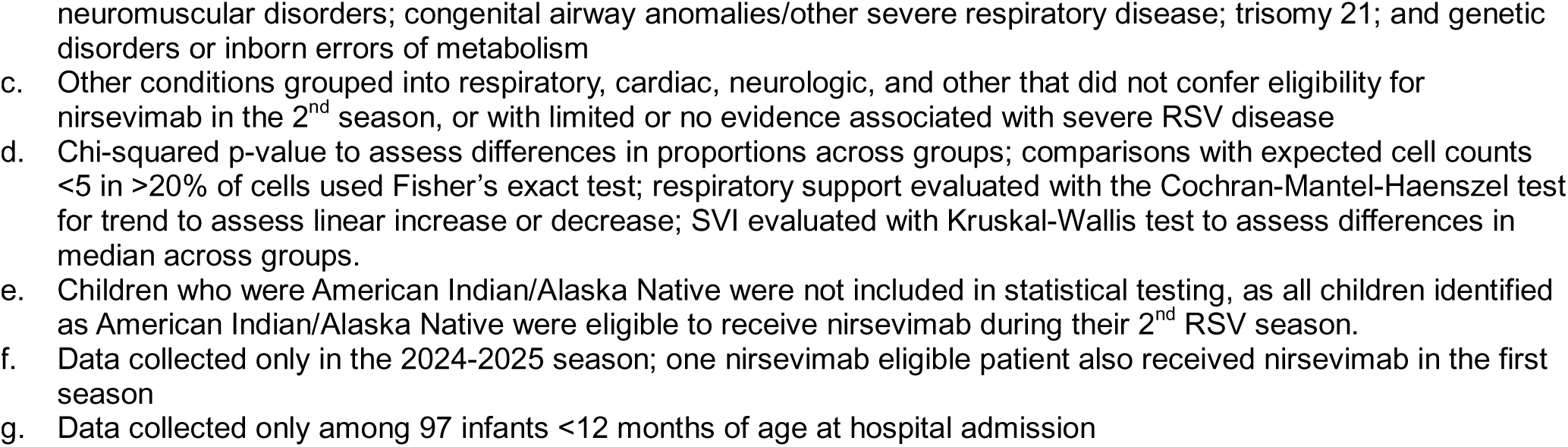
Demographic characteristics by mutually exclusive groups of eligibility status and preexisting health conditions among 574 children aged 8 months to <24 months admitted to the intensive care unit for respiratory syncytial virus (RSV) lower respiratory tract infection in their second RSV season between October 30, 2023 and April 15, 2025 – 30 hospitals, United States.

| Characteristic | Preexisting Health Conditions |  |  |  | P-value<sup>d</sup> |
| --- | --- | --- | --- | --- | --- |
|  | U.S. Nirsevimab eligible criteria n (%)<sup>a</sup> | Other Identified RSV risk conditions n (%)<sup>b</sup> | Other conditions n (%)<sup>c</sup> | None n (%) |  |
| **Total (row %)** | 47 (8.2) | 76 (13.2) | 96 (16.7) | 355 (61.8) | . |
| **RSV season** |  |  |  |  |  |
| 2023–24 | 22 (46.8) | 45 (59.2) | 49 (51.0) | 160 (45.1) | 0.143 |
| 2024–25 | 25 (53.2) | 31 (40.8) | 47 (49.0) | 195 (54.9) |  |
| **Age group at admission** |  |  |  |  |  |
| 8 to <12 months | 10 (21.3) | 15 (19.7) | 11 (11.5) | 61 (17.2) | 0.466 |
| 12 to <16 months | 18 (38.3) | 26 (34.2) | 46 (47.9) | 156 (43.9) |  |
| 16 to <24 months | 19 (40.4) | 35 (46.1) | 39 (40.6) | 138 (38.9) |  |
| **Sex** |  |  |  |  |  |
| Male | 29 (61.7) | 39 (51.3) | 55 (57.3) | 194 (54.6) | 0.687 |
| Female | 18 (38.3) | 37 (48.7) | 41 (42.7) | 161 (45.4) |  |
| **Race and ethnicity<sup>e</sup>** |  |  |  |  |  |
| American Indian/Alaska Native | 6 (12.8) | N/A | N/A | N/A |  |
| White, non-Hispanic | 22 (46.8) | 35 (46.1) | 42 (43.8) | 171 (48.2) | 0.215 |
| Black, non-Hispanic | 12 (25.5) | 17 (22.4) | 22 (22.9) | 58 (16.3) |  |
| Asian, non-Hispanic | 0 | 1 (1.3) | 3 (3.1) | 17 (4.8) |  |
| Hispanic/Latino | 1 (2.1) | 13 (17.1) | 14 (14.6) | 63 (17.7) |  |
| Multiple/Other/Unknown | 6 (12.8) | 10 (13.2) | 15 (15.6) | 46 (13.0) |  |
| **SVI score, median (IQR)** | 46.4 (31.9, 72.3) | 53.5 (34.1, 72.6) | 48.8 (31.9, 68.8) | 45.3 (27.8, 66.5) | 0.398 |
| **Second season nirsevimab received (n=298)<sup>f</sup>** | 3/25 (12.0) | 4/31 (12.9) | 1/47 (2.1) | 0/195 | <.001 |
| **Gestational age (n=97)<sup>g</sup>** | n=10 | n=15 | n=11 | n=61 |  |
| Term (≥37 weeks gestation) | 2 (20.0) | 8 (53.3) | 8 (72.7) | 50 (82.0) | <.001 |
| Preterm (<37 weeks gestation) | 8 (80.0) | 7 (46.7) | 3 (27.3) | 11 (18.0) |  |
| **Respiratory support** |  |  |  |  |  |
| High flow nasal cannula | 14 (29.8) | 25 (32.9) | 40 (41.7) | 159 (44.8) | <.001 |
| Noninvasive ventilation | 14 (29.8) | 38 (50.0) | 46 (47.9) | 175 (49.3) |  |
| Invasive mechanical ventilation | 19 (40.4) | 13 (17.1) | 10 (10.4) | 21 (5.9) |  |
| **Length of hospital stay** | 8.5 (5.0, 11.0) | 8.5 (5.0, 11.0) | 5.0 (3.5, 6.0) | 5.0 (4.0, 6.0) | <.001 |
| **Adverse outcomes** |  |  |  |  |  |
| Extracorporeal membrane oxygenation | 0 | 0 | 2 (2.1) | 0 | N/A |
| In-hospital death | 1 (2.1) | 2 (2.6) | 0 | 0 | N/A |
| Vasoactive infusions | 8 (17.0) | 10 (13.2) | 6 (6.3) | 9 (2.5) | <.001 |

Abbreviations: IQR – Interquartile range; SVI – Social vulnerability index
- Chronic lung disease (CLD) of prematurity requiring medical support (chronic corticosteroid therapy, diuretic therapy, or supplemental oxygen) during the 6-month period before the start of the second RSV season, 2) severe immunocompromise, 3) cystic fibrosis with either manifestations of severe lung disease or weight-for-length <10th percentile, or 4) being American Indian/Alaska Native - Congenital heart disease with history of cardiac repair, pulmonary hypertension, or congestive heart failure;
neuromuscular disorders; congenital airway anomalies/other severe respiratory disease; trisomy 21; and genetic disorders or inborn errors of metabolism
- c. Other conditions grouped into respiratory, cardiac, neurologic, and other that did not confer eligibility for nirsevimab in the 2<sup>nd</sup> season, or with limited or no evidence associated with severe RSV disease - d. Chi-squared p-value to assess differences in proportions across groups; comparisons with expected cell counts <5 in >20% of cells used Fisher's exact test; respiratory support evaluated with the Cochran-Mantel-Haenszel test for trend to assess linear increase or decrease; SVI evaluated with Kruskal-Wallis test to assess differences in median across groups. - e. Children who were American Indian/Alaska Native were not included in statistical testing, as all children identified as American Indian/Alaska Native were eligible to receive nirsevimab during their 2<sup>nd</sup> RSV season. - f. Data collected only in the 2024-2025 season; one nirsevimab eligible patient also received nirsevimab in the first season - g. Data collected only among 97 infants <12 months of age at hospital admission

These conditions were: hemodynamically significant cardiac disease^15–17^; congenital airway anomalies and other chronic lung disease^15,17^; neuromuscular disease or neurologic conditions^15,17^; trisomy 21^16,17^; and genetic disorders or inborn errors of metabolism.^15,17^ Other conditions not included in U.S. eligibility criteria or not known to be associated with increased risk for severe RSV disease were included as other conditions. Supplemental Table 1 describes which conditions were collected on the Overcoming RSV case report form that align with those conditions. For example, for this investigation, hemodynamically significant cardiac disease was comprised of the following from the case report form: congenital heart disease with history of cardiac repair, moderate/severe pulmonary hypertension, and congestive heart failure requiring medication. Prematurity is also considered among other identified RSV risk conditions by some countries; in our investigation data on prematurity was collected only for children <12 months of age at time of hospitalization. Any clinical viral or bacterial testing results available in the medical record were abstracted. Receipt of nirsevimab was collected only during study year 2, the 2024−25 RSV season.

Information on demographic characteristics was described and compared by preexisting health conditions groups. Chi-squared and Kruskal-Wallis tests were used to identify statistically significant differences in demographic characteristics by eligibility status and preexisting health conditions group. We additionally reported the highest level of respiratory support received by preexisting health conditions groups, comparing differences in the distribution of respiratory support and condition status group using the Cochran-Mantel-Haenszel (CMH) test for trend.

All data analyses were conducted using SAS (version 9.4, SAS Institute). This activity was reviewed by the Centers for Disease Control and Prevention, deemed not research, and was conducted consistent with applicable federal law and CDC policy (45 C.F.R. part 46.102(l)(2), 21 C.F.R. part 56; 42 U.S.C. Sect. 241(d); 5 U.S.C. Sect. 552a; 44 U.S.C. Sect. 3501 et seq.).

## Results

Among 574 children experiencing severe RSV and PICU admission during their second RSV season, 47 (8.2%) were in the second season U.S. nirsevimab eligible criteria group (Table 1). Additionally, 76 children (13.2%) had other identified RSV risk conditions, 96 children (16.7%) had other conditions, and most (61.8%) had no conditions (Table 2). Across preexisting health conditions groups, children were similar by season of admission, race and ethnicity, age at admission, sex, and social vulnerability index (SVI). Among infants <12 months of age (n=97), preterm birth was more common among those who were eligible for nirsevimab in their second season (Table 1).

**Table 2.**
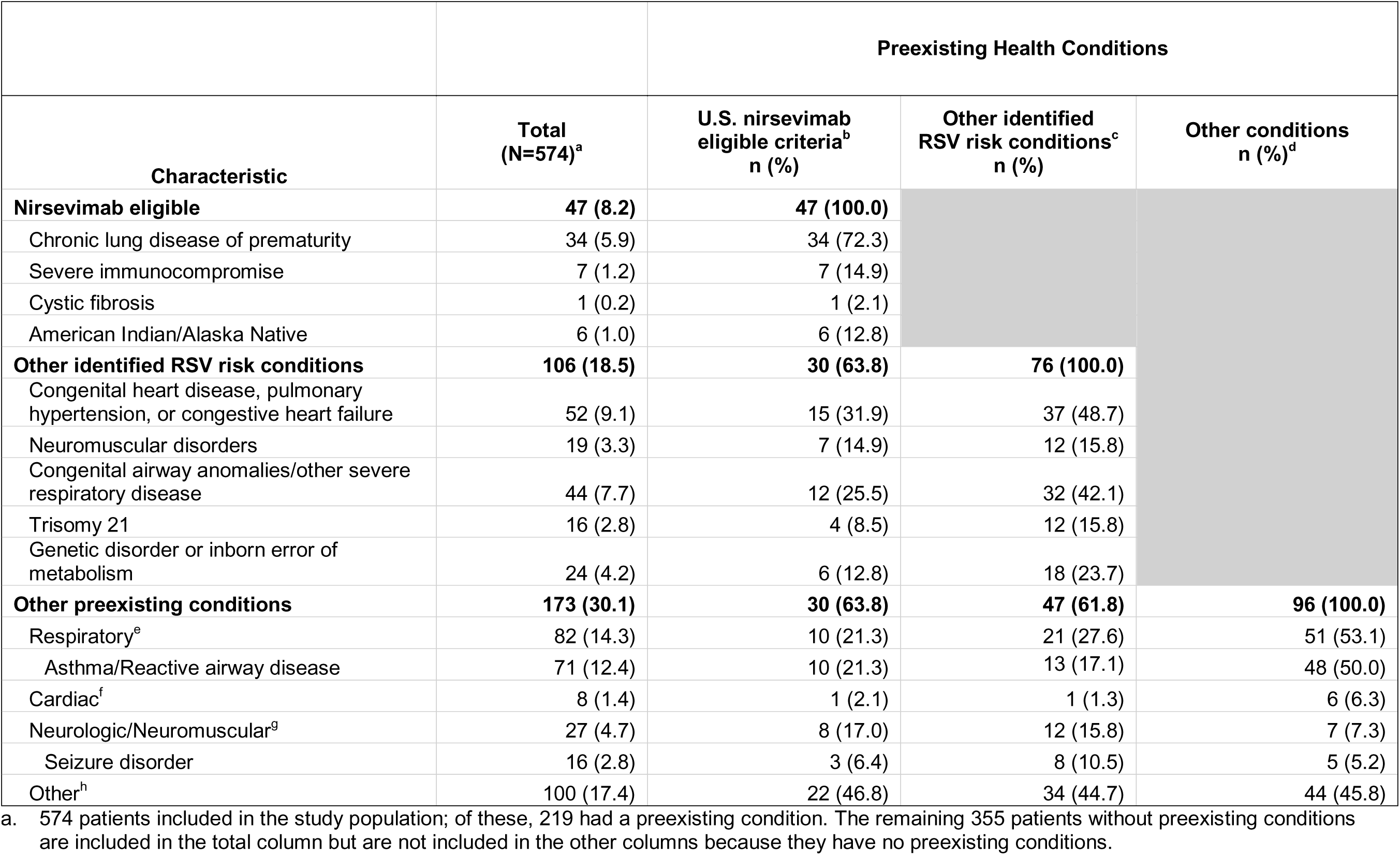
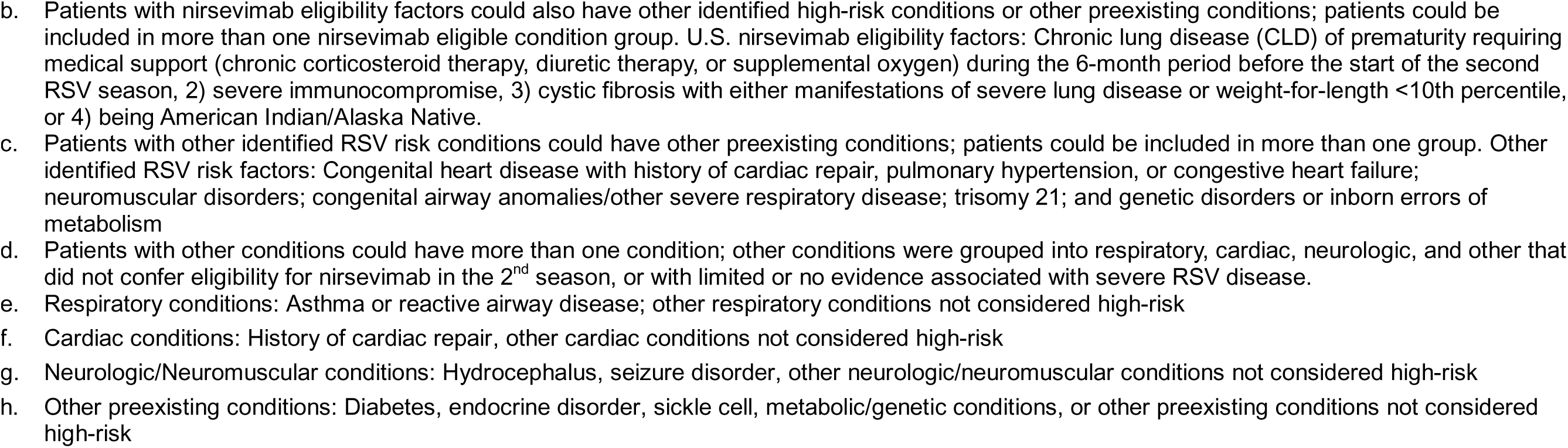
Distribution of health conditions among children aged 8 months to <24 months admitted to the intensive care unit for respiratory syncytial virus (RSV) lower respiratory tract infection in their second RSV season by mutually exclusive groups of eligibility status and preexisting health conditions between October 30, 2023 and April 15, 2025 - 30 hospitals, United States.

|  |  | Preexisting Health Conditions |  |  |
| --- | --- | --- | --- | --- |
| Characteristic | Total<br>(N=574)<sup>a</sup> | U.S. nirsevimab<br>eligible criteria<sup>b</sup><br>n (%) | Other identified<br>RSV risk conditions<sup>c</sup><br>n (%) | Other conditions<br>n (%)<sup>d</sup> |
| **Nirsevimab eligible** | **47 (8.2)** | **47 (100.0)** |  |  |
| Chronic lung disease of prematurity | 34 (5.9) | 34 (72.3) |  |  |
| Severe immunocompromise | 7 (1.2) | 7 (14.9) |  |  |
| Cystic fibrosis | 1 (0.2) | 1 (2.1) |  |  |
| American Indian/Alaska Native | 6 (1.0) | 6 (12.8) |  |  |
| **Other identified RSV risk conditions** | **106 (18.5)** | **30 (63.8)** | **76 (100.0)** |  |
| Congenital heart disease, pulmonary hypertension, or congestive heart failure | 52 (9.1) | 15 (31.9) | 37 (48.7) |  |
| Neuromuscular disorders | 19 (3.3) | 7 (14.9) | 12 (15.8) |  |
| Congenital airway anomalies/other severe respiratory disease | 44 (7.7) | 12 (25.5) | 32 (42.1) |  |
| Trisomy 21 | 16 (2.8) | 4 (8.5) | 12 (15.8) |  |
| Genetic disorder or inborn error of metabolism | 24 (4.2) | 6 (12.8) | 18 (23.7) |  |
| **Other preexisting conditions** | **173 (30.1)** | **30 (63.8)** | **47 (61.8)** | **96 (100.0)** |
| Respiratory<sup>e</sup> | 82 (14.3) | 10 (21.3) | 21 (27.6) | 51 (53.1) |
| Asthma/Reactive airway disease | 71 (12.4) | 10 (21.3) | 13 (17.1) | 48 (50.0) |
| Cardiac<sup>f</sup> | 8 (1.4) | 1 (2.1) | 1 (1.3) | 6 (6.3) |
| Neurologic/Neuromuscular<sup>g</sup> | 27 (4.7) | 8 (17.0) | 12 (15.8) | 7 (7.3) |
| Seizure disorder | 16 (2.8) | 3 (6.4) | 8 (10.5) | 5 (5.2) |
| Other<sup>h</sup> | 100 (17.4) | 22 (46.8) | 34 (44.7) | 44 (45.8) |

a. 574 patients included in the study population; of these, 219 had a preexisting condition. The remaining 355 patients without preexisting conditions are included in the total column but are not included in the other columns because they have no preexisting conditions.

Nirsevimab receipt was not collected in 2023–24, and among children admitted in the 2024–25 season, when receipt of nirsevimab was collected (n=298), eight children (2.7%) received nirsevimab in their second season (Table 1, Supp. Table 2). Three of these eight children were classified in the U.S. nirsevimab eligible criteria group, and one U.S. nirsevimab eligible child also received nirsevimab in the first RSV season. The five children not in the U.S. nirsevimab eligible criteria group included four children with other identified RSV risk conditions (3 congenital heart disease with history of cardiac repair, including one child with trisomy 21, and one with neuromuscular weakness), and one in the other condition group (history of cardiac repair unspecified).

Among the 47 U.S. nirsevimab eligible criteria group patients, 34 (72.3%) had CLD requiring medical support within 6 months preceding the RSV season, one (2.1%) had cystic fibrosis with manifestations of severe lung disease, 6 (12.8%) were American Indian/Alaska Native, and 7 (14.9%) had severe immunocompromise (including one who also had CLD) (Table 2). Among children not in the U.S. nirsevimab eligible criteria group (n=527), 76 (14.4%) had other identified RSV risk conditions, 96 (18.2%) had other health conditions, and 355 (67.4%) had no preexisting conditions. In children with other identified RSV risk conditions, 37 (48.7%) had congenital heart disease, pulmonary hypertension, or congestive heart failure, 32 (42.1%) had congenital airway anomalies or severe respiratory disease, 12 (15.8%) had trisomy 21, and 18 (23.7%) had genetic disorders or inborn errors of metabolism. Children with other conditions who experienced severe RSV disease (n=96) most commonly had respiratory disorders (n=51, 53.1%).

The proportion of children requiring invasive mechanical ventilation increased significantly across preexisting health conditions groups from no conditions to U.S. nirsevimab eligible criteria (p_trend_ <0.001, Figure 1). Among 47 children with nirsevimab eligible criteria, 19 (40.4%) required invasive mechanical ventilation (Table 1, Figure 1). In contrast, invasive mechanical ventilation was required for 17.1% of those with other identified RSV risk conditions, 10.4% of those with other conditions, and 5.9% of those without conditions (Figure 1).

**Figure 1.**
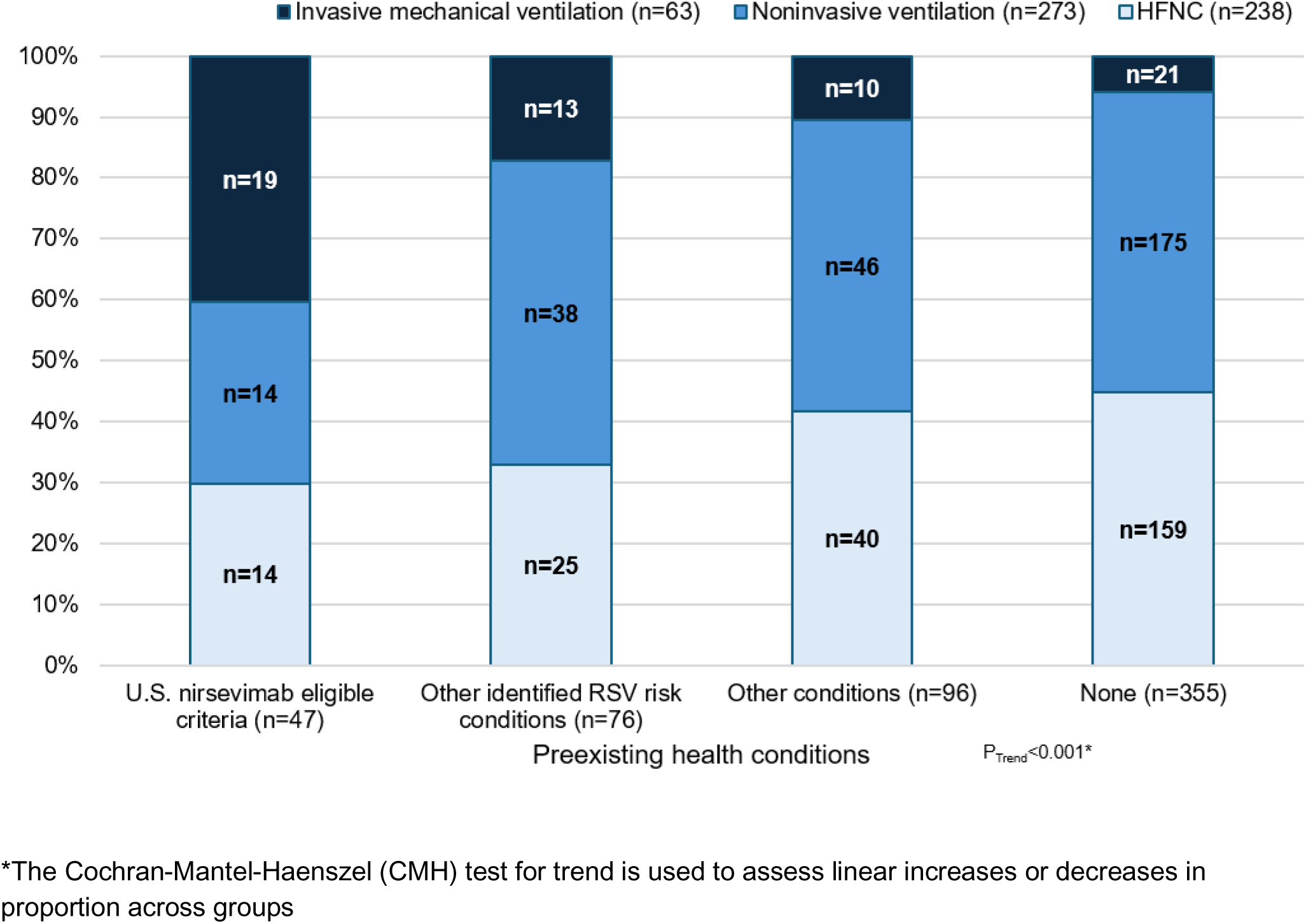
Proportion of children in each preexisting health conditions group whose highest level of respiratory support during the pediatric intensive care unit stay was invasive mechanical ventilation, noninvasive ventilation, or high-flow nasal cannula (HFNC). Inclusion criteria required that all children receive high flow nasal cannula or greater support.

Of the two children who required extracorporeal membrane oxygenation (ECMO) support, both had other preexisting conditions. One had a history of seizure disorder and the other had a history of asthma/reactive airway disease; both survived (Supp. Table 3). Neither had other viruses detected but both had bacterial coinfections. Three children died, including one in the U.S. nirsevimab eligible criteria group who had a history of bronchopulmonary dysplasia, immune suppression, an upper airway disorder, and a chromosomal disorder; no viral or bacterial co-infections were identified, and cause of death for this patient was attributed to multiorgan failure due to post-transplantation complications. The other two patients had other identified RSV risk conditions: one patient with a chromosomal disorder who also had viral codetection with influenza and adenovirus, and cause of death noted to be severe lung disease, and one with a neurologic condition, no co-infections, and cause of death of severe neurologic injury (Supp. Table 3).

## Discussion

This multicenter cohort investigation included 574 U.S. children aged 8–<24 months old admitted to the PICU for severe RSV during their second RSV season. We found that approximately one-fifth of the children were in the defined groups that either met U.S. nirsevimab eligibility criteria (8%) or that had an identified RSV risk condition associated with known risk for severe RSV (13%). Approximately 17% had other preexisting conditions, while the majority (62%) of children did not have a preexisting health condition.

The approximately 8% of children in this cohort who met U.S. nirsevimab eligibility criteria for their second season comprised a disproportionately higher percentage than the prevalence of this group in the U.S. population, which has been previously estimated as occurring in 2%–4% of U.S. children.^19^ This previously cited prevalence was likely an overestimate because it does not account for the complex phenotypes incorporated into the preexisting health conditions that currently comprise recommended high-risk U.S. nirsevimab eligibility criteria.^5,19^ For example, U.S. criteria for nirsevimab eligibility for chronic lung disease of prematurity requires medical support in the six month period before the start of the second RSV season; this requirement is not reflected in overall U.S. prevalence reported.

Among this entire PICU cohort, preexisting identified RSV risk conditions also appear to be overrepresented compared to estimated prevalence in the U.S. population. Approximately 9% of children in this cohort had congenital heart disease, moderate-to-severe pulmonary hypertension, or congestive heart failure. Although it is difficult to estimate prevalence in the general population of U.S. children, the combined prevalence of these conditions is likely to be no more than 1%-2%.^19–22^ Chronic respiratory conditions including congenital airway anomalies and other severe respiratory disease were present in nearly 8% of this PICU cohort; although direct population comparators are limited for this diverse category, congenital airway anomalies are estimated to affect <0.01% of live births.^23^ Neuromuscular disorders, including muscular dystrophy, spastic quadriplegia, static encephalopathy, and neuromuscular weakness, were present in approximately 3% of patients, but the population prevalence of these individual disorders ranges from <0.01% to 0.3%.^24,25^ Trisomy 21 comprised nearly 3% of this cohort, whereas the U.S. prevalence estimate of trisomy 21 is 0.2%.^26^

Overall, these previously identified RSV risk conditions that are not included in U.S. nirsevimab eligibility criteria are rare to uncommon in the general U.S. population. Given their potentially increased prevalence among young children admitted to the PICU with severe RSV disease, this may suggest an increased risk of severe disease in children with these conditions and warrants further investigation.^27^ Changes to RSV immunization recommendations are not without precedent, as palivizumab recommendations changed after initial approval as more information was gathered about those most at risk and cost effectiveness of the product.^28^ Findings from this investigation and additional studies could inform evidence on the condition-specific risk of severe RSV outcomes for organizations considering RSV immunization policies.

Outside of the U.S., guidance for second season nirsevimab immunization varies. Many countries recommend nirsevimab for children with hemodynamically significant congenital heart disease.^29–35^ Canada recommends nirsevimab specifically for airway anomalies that impair clearance of secretions,^36^ and the UK indicates nirsevimab if these abnormalities result in the receipt of long-term ventilation,^37^ but Spain, Chile, and South Korea recommend nirsevimab for children with these airway anomalies regardless of their need for respiratory support.^31,32,35^ Paraguay and South Korea recommend nirsevimab with hemodynamically significant congenital heart disease.^35,38^ In the present, U.S.-based analysis, over half of the children who received nirsevimab did not qualify for nirsevimab receipt, suggesting a healthcare provider considered them of sufficiently high risk to receive the immunization. In this analysis, the rate of invasive mechanical ventilation among those with identified RSV risk conditions that are not are not included in U.S. nirsevimab eligibility criteria were elevated compared to those with other and no preexisting conditions. Additionally, severe outcomes did occur among those with other preexisting conditions, further underscoring the need for more evidence about health conditions that increase risk for severe RSV disease in a child’s second RSV season.

### Limitations

This analysis is subject to multiple limitations. First, the proportion of children who were eligible to receive nirsevimab during their second RSV season was small, limiting sample size for statistical comparisons between preexisting health conditions groups. Second, clinical data were abstracted from records at the study site hospitals, and medical information from any referring institution as well as complete preexisting health information may not have been available. Third, site-level variation in clinical testing practices and admission to the PICU could impact the representativeness of these results. Fourth, information on nirsevimab receipt was collected only for the 2024−25 RSV season; however, while this information was not collected for children hospitalized during the 2023−24 season, nirsevimab availability was very limited in this first season after its recommendation.^39^ Fifth, information on premature status was collected only for children <12 months of age; therefore, we could not separately assess prematurity as a risk factor in children 12 months or older. Sixth, details about the health consequences and severity of each preexisting condition were limited. For example, the hemodynamic significance of cardiac conditions could not be assessed. Finally, as this analysis was conducted only among children admitted to the PICU, we could not estimate incidence of PICU admission or other outcomes among children with these conditions.

## Conclusions

Among 574 children aged 8–23 months admitted to the PICU for severe RSV disease in their second RSV season, the majority did not have a preexisting medical condition and fewer than 10% had risk factors that made them eligible to receive nirsevimab. We identified additional children (13.2%) who had other identified RSV risk conditions; the majority were severe cardiac and respiratory conditions but also included neuromuscular and genetic disorders. These findings could inform potential considerations for expanding nirsevimab immunization eligibility among additional high-risk children in their second RSV season in the United States. Further study is needed on these and other potential health conditions that may be risk factors for severe RSV disease during a child’s second RSV season, on the severity of clinical outcomes in this second season population, and on the effectiveness of nirsevimab in preventing acute respiratory failure in children at high risk for severe RSV disease.

## Data Availability

Data are not publicly available. Deidentified participant data in a limited dataset may be made available upon formal request for a specified purpose for researchers whose proposed use of the data has been approved.

## Funding/Acknowledgements

This work was funded by CDC through contract # 75D30122C13330 to Boston Children’s Hospital.

## Disclaimer

The findings and conclusions in this report are those of the authors and do not necessarily represent the official position of the Centers for Disease Control and Prevention.

## Author Contributions

Drs. Simeone and Zambrano had full access to all of the data in the study and take responsibility for the integrity of the data and the accuracy of the data analysis.

*Concept and Design*: Simeone, Zambrano, Campbell, Randolph, Newhams, Orzel-Lockwood

*Acquisition, analysis, or interpretation of data*: All authors

*Drafting of the manuscript*: Simeone and Zambrano

*Critical review of the manuscript*: All authors

*Statistical analysis*: Simeone and Zambrano

*Supervision:* Campbell and Randolph

## Overcoming RSV Investigators

**Alabama:** <u>Children’s of Alabama, Birmingham.</u> Michele Kong, Meghan Murdock, Heather Kelley; Candice Colston; **Arkansas:** <u>Arkansas Children’s Hospital, Little Rock.</u> Katherine Irby, Ronald C. Sanders, Masson Yates, Ashlyn Madding; **California:** <u>Rady Children’s Hospital, San Diego.</u> Melissa A. Cameron, Michael Henne; **California:** <u>UCSF Benioff Children’s Hospital, San Francisco.</u> Matt S. Zinter, Mary Prahl; **Colorado:** <u>Children’s Hospital Colorado, Aurora.</u> Aline B. Maddux, Ariana Valenzuela, Natasha Baig, Lexi Petruccelli, Rachel Greer, Heidi Sauceda, Sara Shankman, Lanae Dailey, Jaime Laurin; **Delaware:** <u>Nemours Children’s Hospital, Wilmington, Delaware; Sidney Kimmel Medical College, Thomas Jefferson University, Philadelphia, PA.</u> Jigar C. Chauhan, Scott L. Weiss, Rebecca Clifford, Jenna Lapira; **Georgia:** <u>Emory University School of Medicine and Children’s Healthcare of Atlanta, Atlanta.</u> Satoshi Kamidani, Mark D. Gonzalez, Caroline R. Ciric, Jong-Ha C. Choi, Gabriella Ess, Anna K. Mitchell, Luis W. Salazar, Janelle J. Spencer-Ramirez; **Illinois:** <u>Ann & Robert H. Lurie Children’s Hospital of Chicago, Chicago.</u> Kelly N. Michelson, Bria M. Coates, Sarah Jae Wershil; **Indiana:** <u>Riley Hospital for Children, Indianapolis.</u> Samina S. Bhumbra, Mary Stumpf; Iowa: <u>Stead Family Children’s Hospital, Iowa City.</u> Kari Wellnitz; **Louisiana:** <u>Children’s Hospital of New Orleans, New Orleans.</u> Tamara T. Bradford; **Massachusetts:** <u>Boston Children’s Hospital, Boston.</u> Adrienne G. Randolph, Margaret M. Newhams, Amber O. Orzel-Lockwood, Jemima M. Calixte, Amira Toivonen, Callum Ackerman; **Minnesota:** <u>University of Minnesota Masonic Children’s Hospital, Minneapolis,</u> Janet R. Hume, Judy Kyrkos, Joanna Ly, Katrina Saladin; **Minnesota:** <u>Mayo Clinic, Rochester.</u> Emily R. Levy, Chloe A. Schluttner, Carly R. Crowley; **Mississippi:** <u>Children’s Hospital of Mississippi, Jackson.</u> Lora Martin, Joseph M. Majure, Lacy Malloch, Maygan Martin; **Missouri:** <u>Children’s Mercy Kansas City, Kansas City.</u> Jennifer E. Schuster, Berenice Marrufo, Allison Scott; **New Jersey:** <u>Cooperman Barnabas Medical Center, Livingston.</u> Shira J. Gertz, Elizabeth Ricciardi; **New York:** <u>Bernard & Millie Duker Children’s Hospital, Albany.</u> Saul R. Hymes, Ilana Harwayne-Gidansky; **New York:** <u>Columbia University Irving Medical Center, New York City.</u> Melissa S. Stockwell, Celibell Y. Vargas, Raul A. Silverio Francisco, Thomas J. Connors; **North Carolina:** <u>University of North Carolina at Chapel Hill, Chapel Hill.</u> Stephanie P. Schwartz, Tracie C. Walker, Alison Mathers; **Ohio:** <u>Akron Children’s Hospital, Akron.</u> Ryan A. Nofziger, Christopher Page-Goertz, Nicole Twinem, Olivia Kennedy; **Ohio:** <u>Cincinnati Children’s Hospital, Cincinnati.</u> Mary Allen Staat, Chelsea C. Rohlfs; **Ohio:** <u>UH Rainbow Babies and Children’s Hospital, Cleveland.</u> Steven Shein, Kenneth Remy, Rajashri Rasal; **Oregon:** <u>Oregon Health and Science University, Portland.</u> Judith A. Guzman-Cottrill, Chelsea Heisler; **Pennsylvania:** <u>Children’s Hospital of Philadelphia, Philadelphia.</u> Kathleen Chiotos, Alanah Mckelvey, Jenny Bush, Rebecca Douglas, Myooran Sivarupan, Glory Edioma, Taylor A. Slocumb; **South Carolina:** <u>MUSC Children’s Health, Charleston.</u> Austin Biggs, Zach Rusler; **Tennessee:** <u>Monroe Carell Jr. Children’s Hospital at Vanderbilt, Nashville.</u> Natasha B. Halasa, Laura S. Stewart, Kristina A. Betters, Annika Conlee; Madeline Spradley; Adriana Blanco Vasquez, Eliana Rebolledo; **Texas:** <u>University of Texas Southwestern, Children’s Medical Center Dallas, Dallas.</u> Mia Maamari, Cindy Bowens; **Utah:** <u>University of Utah and Primary Children’s Hospital, Salt Lake City.</u> Hillary Crandall; **Washington:** <u>Seattle Children’s Hospital, Seattle.</u> Danielle Zerr, Amanda Adler

---

**Supp. Table 1.** U.S. and other country-specific recommendations for administration of nirsevimab during the second RSV season.*

| Condition | Countries outside U.S. where included in second season nirsevimab eligibility | Conditions specified and collected on the *Overcoming RSV* case report form that align with condition | Notes |
| --- | --- | --- | --- |
| **U.S. criteria for nirsevimab eligibility** |  |  |  |
| Chronic lung disease of prematurity requiring ongoing medical support in the 6 months before hospitalization | US<sup>1</sup>, CA<sup>2</sup>, AU<sup>3</sup>, SP<sup>4</sup>, CL<sup>5</sup>, FR<sup>6</sup>, JP<sup>7,8</sup>, UK<sup>9</sup>, KR<sup>10</sup> | Bronchopulmonary dysplasia, with receipt of medical support (chronic corticosteroid therapy, diuretic therapy, or supplemental oxygen) in the past 6 months | Medical support is defined as chronic corticosteroid therapy, diuretic therapy, or supplemental oxygen. Australia uses a definition that considers oxygen support beyond 36 weeks gestation or at hospital discharge. In Spain, patients with bronchopulmonary dysplasia, especially those with severe BPD, with priority given to those needing treatment in the six months before season onset. In Chile, medical support is not required. In the UK, only children with moderate or severe chronic lung disease who were preterm who had compatible x-ray changes and who continue to receive supplemental oxygen or respiratory support at 36 weeks gestational age considered eligible. |
| Cystic fibrosis | US<sup>1</sup>, CA<sup>2</sup>, AU<sup>3</sup>, SP<sup>4</sup>, CL<sup>5</sup>, KR<sup>10</sup> | Cystic fibrosis, with history of hospitalization in the first year of life for a CF pulmonary exacerbation OR persistent abnormalities on chest x-ray or chest CT that persist even when stable and not having pulmonary exacerbations OR weight-for-length <10<sup>th</sup> percentile | The U.S. and most other included countries only consider cystic fibrosis patients as eligible if they have manifestations of severe lung disease or weight-for-length percentile <10<sup>th</sup> percentile. Weight-for-height percentiles calculated were calculated for each patient in this analysis using the World Health Organization's Child Growth Standards for children <2 years of age (https://www.who.int/toolkits/child-growth-standards/standards). |
| Severe immunocompromise/immunodeficiency | US<sup>1</sup>, CA<sup>2</sup>, AU<sup>3</sup>, SP<sup>4</sup>, CL<sup>5</sup>, JP<sup>7,8</sup>, UK<sup>9</sup>, KR<sup>10</sup> | Any immunosuppressive or oncologic disorder | Australia provides malignancy, solid organ transplant, hematopoietic stem cell transplant, or inborn errors of immunity as examples. The United Kingdom specifies severe combined immunodeficiency syndrome, or any condition that renders a child unable to mount a T-cell response or produce antibodies against infectious agents. |
|  |  |  | Spain specifies hematologic malignancies, primary immunodeficiency, or treatment with immunosuppressive medications. France identifies immunosuppression as a high-risk underlying condition but does not specifically indicate nirsevimab during the second season for this reason. |
| American Indian/Alaska Native or other indigenous population | US<sup>1</sup> | Race/ethnicity specified as American Indian or Alaska Native | The U.S. specifies American Indian or Alaskan Native, but we considered mention of any other indigenous populations when assessing other country criteria. |
| **Other identified RSV risk health conditions sufficient for second season nirsevimab recommendation outside the U.S.** |  |  |  |
| Hemodynamically significant cardiac disease | CA<sup>2</sup>, AU<sup>3</sup>, SP<sup>4</sup>, CL<sup>5</sup>, FR<sup>6</sup>, JP<sup>7,8</sup>, PY<sup>11</sup>, KR<sup>10</sup> | Congenital heart disease with history of cardiac repair, moderate/severe pulmonary hypertension, congestive heart failure requiring medication | Chile specifies hemodynamically significant congenital heart disease (CHD) not resolved or cyanotic CHD secondary to high-complexity CHD. The UK recognizes CHD as a high-risk underlying condition but does not specifically recommend that children with hemodynamically significant CHD receive nirsevimab in their second RSV season. Spain specifies history of cardiac bypass. France identifies children with history of cardiac shunt as being a vulnerable population, but there is no explicit recommendation for second season nirsevimab for this group. |
| Severe congenital airway anomalies/pulmonary malformations/other severe respiratory disease | CA<sup>2</sup>, SP<sup>4,5</sup>, CL<sup>5,12</sup>, UK<sup>9</sup>, KR<sup>10</sup> | Chronic lung disease (other than U.S. nirsevimab eligibility factors), assistance required for clearance secretion, or any upper airway disorder (e.g. tracheomalacia/laryngomalacia, congenital airway disorders, obstructive sleep apnea) | Canada and South Korea indicate nirsevimab if these airway anomalies impair clearance of secretions. UK indicates nirsevimab in the second season of these abnormalities have resulted in receipt of long-term ventilation. |
| Neuromuscular disease or neurological conditions | CA<sup>2</sup>, AU<sup>3</sup>, SP<sup>4</sup>, CL<sup>5,12</sup>, KR<sup>10</sup> | Muscular dystrophy, static encephalopathy, spastic quadriplegia, or neuromuscular weakness | Canada and South Korea specify neuromuscular disease impairing clearance of secretions. Australia specifies neuromuscular disease that impairs respiratory function, including congenital, hereditary, or degenerative central nervous system disorders, cerebral palsy, brain or spinal cord conditions affecting respiratory control/function, neuromuscular disorders, and conditions with impaired swallowing/coughing or with aspiration risk. France identifies children with neuromuscular conditions impacting respiratory function as a vulnerable group, but there is no explicit second season recommendation for nirsevimab administration in this group. |
| Trisomy 21 | AU<sup>3</sup>, SP<sup>4</sup>, CL<sup>5,12</sup>, JP<sup>7,8</sup>, KR<sup>10</sup> | Trisomy 21, presence of congenital heart disease not required | France identifies children with trisomy 21 with complications affecting respiratory function as a vulnerable group but does not explicitly recommend nirsevimab during the second RSV season for this group. |
| Prematurity, with or without chronic lung disease | AU<sup>3</sup>, SP<sup>4</sup>, CL<sup>5,12</sup> JP<sup>7,8</sup> | Collected only for children <12 months of age at hospitalization. | Gestational age cutoff for 2<sup>nd</sup> season administration among children born prematurely varies by country, including Australia (<32 weeks), Spain (<35 weeks for infants <12 months of age), Chile (<32 weeks or birthweight <1500g), Japan (<29 weeks for infants <12 months of age). |

Abbreviations: US = United States; CA = Canada; AU = Australia; SP = Spain; CL = Chile; FR = France; JP = Japan; PY = Paraguay; UK = United Kingdom, KR = South Korea
\* As of February 12, 2026; information herein obtained from review of published recommendations in PubMed and results of internet searches citing recommendations from other country National Immunization Technical Advisory Groups.

**Supp. Table 2.**
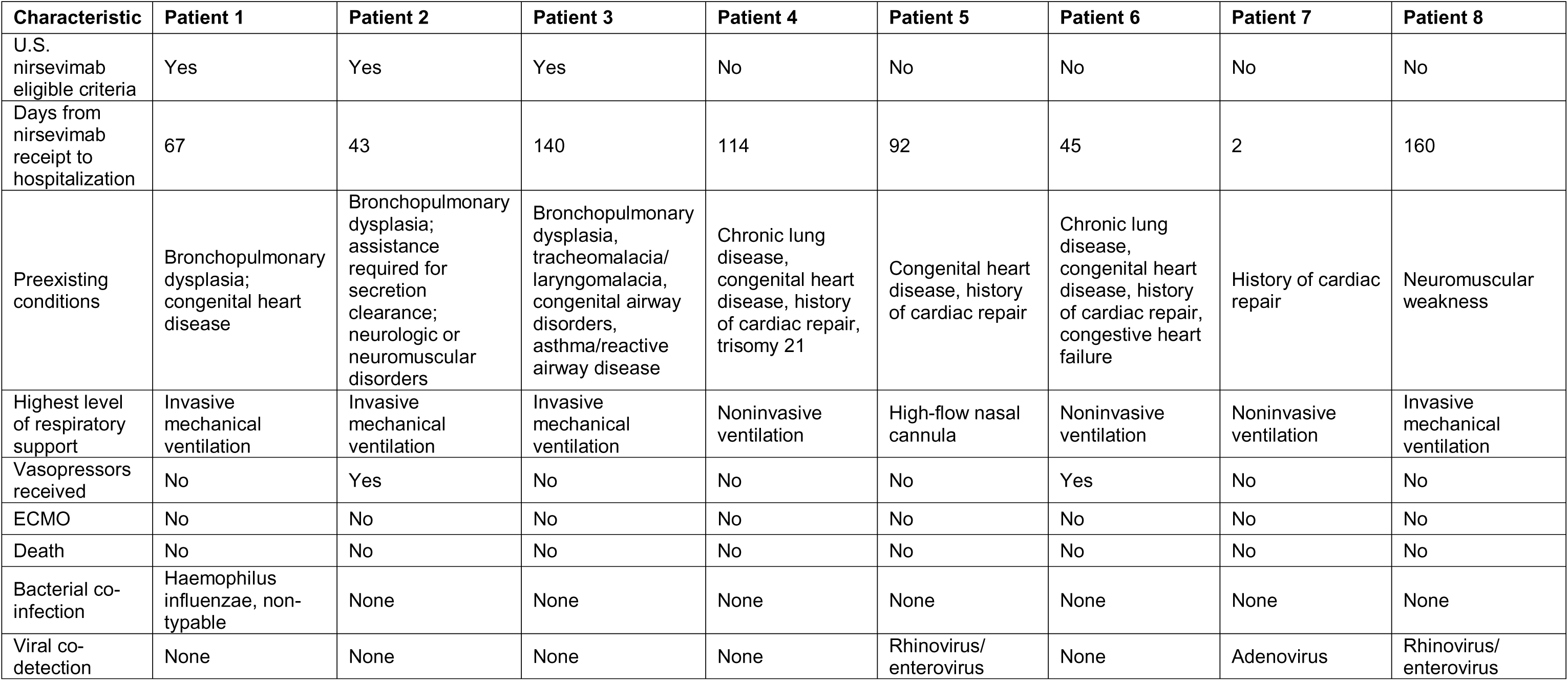
Clinical characteristics of children aged 8 months to <24 months admitted to the intensive care unit for respiratory syncytial virus (RSV) lower respiratory tract infection in their second RSV season who received nirsevimab during their second RSV season.

| Characteristic | Patient 1 | Patient 2 | Patient 3 | Patient 4 | Patient 5 | Patient 6 | Patient 7 | Patient 8 |
| --- | --- | --- | --- | --- | --- | --- | --- | --- |
| U.S. nirsevimab eligible criteria | Yes | Yes | Yes | No | No | No | No | No |
| Days from nirsevimab receipt to hospitalization | 67 | 43 | 140 | 114 | 92 | 45 | 2 | 160 |
| Preexisting conditions | Bronchopulmonary dysplasia; congenital heart disease | Bronchopulmonary dysplasia; assistance required for secretion clearance; neurologic or neuromuscular disorders | Bronchopulmonary dysplasia, tracheomalacia/laryngomalacia, congenital airway disorders, asthma/reactive airway disease | Chronic lung disease, congenital heart disease, history of cardiac repair, trisomy 21 | Congenital heart disease, history of cardiac repair | Chronic lung disease, congenital heart disease, history of cardiac repair, congestive heart failure | History of cardiac repair | Neuromuscular weakness |
| Highest level of respiratory support | Invasive mechanical ventilation | Invasive mechanical ventilation | Invasive mechanical ventilation | Noninvasive ventilation | High-flow nasal cannula | Noninvasive ventilation | Noninvasive ventilation | Invasive mechanical ventilation |
| Vasopressors received | No | Yes | No | No | No | Yes | No | No |
| ECMO | No | No | No | No | No | No | No | No |
| Death | No | No | No | No | No | No | No | No |
| Bacterial co-infection | Haemophilus influenzae, non-typable | None | None | None | None | None | None | None |
| Viral co-detection | None | None | None | None | Rhinovirus/enterovirus | None | Adenovirus | Rhinovirus/enterovirus |

**Supp. Table 3.** Clinical characteristics of children aged 8 months to <24 months admitted to the intensive care unit for respiratory syncytial virus (RSV) lower respiratory tract infection in their second RSV season who received ECMO or died during their hospitalization. For privacy, age is rounded to the closest month and gestational age is given as a range.

| Characteristic | Death 1 (no ECMO) | Death 2 (no ECMO) | Death 3 (no ECMO) | ECMO 1 Survivor | ECMO 2 Survivor |
| --- | --- | --- | --- | --- | --- |
| Age (months) | 16 to <24 months | 16 to <24 months | 12 to <16 months | 12 to <16 months | 8 to <12 months |
| Preexisting health conditions groups | U.S. nirsevimab eligible criteria | Other identified RSV risk conditions | Other identified RSV risk conditions | Other conditions | Other conditions |
| Preexisting conditions | Bronchopulmonary dysplasia, immune suppression, unspecified chromosomal disorder | Unspecified chromosomal disorder | Neurologic or neuromuscular condition | Seizure disorder | Asthma/reactive airway disease |
| Highest level of respiratory support | Invasive mechanical ventilation | Invasive mechanical ventilation | Invasive mechanical ventilation | Invasive mechanical ventilation | Invasive mechanical ventilation |
| Vasopressors received | Yes | Yes | Yes | Yes | No |
| Viral co-detection | None | Influenza, adenovirus | None | None | None |
| Bacterial co-infection | None | None | None | *Streptococcus pneumoniae* | *Methicillin-susceptible Staphylococcus aureus*; gram negative rods |
| Patient cause of death | Multiorgan failure due to post-transplantation complications | Severe lung disease | Severe neurologic injury | N/A | N/A |

